# Trait-Level Rather Than Individual-Level Resolution in Fitness Genomics: A Population-Calibrated Variant Prioritization Framework in a South Asian Cohort

**DOI:** 10.64898/2026.11.10.26367795

**Authors:** Kalyan Ram Uppaluri, Hima Jyothi Challa, Krishna Kumar Vempati, Lakshmi Nirmala Kadali, Madhu Bhanu Varma Rayala, Jayanthy Prahashit, Balaji Malluri

**Affiliations:** Uppaluri K&H Personalized Medicine Clinic Private Limited. (GenepoweRx), Hyderabad, India

## Abstract

Translating candidate gene panels into objective physiological trait interpretations remains a major hurdle in exercise genomics. Conventional total genotype scoring suffers from severe panel-composition bias, fails to evaluate unannotated or novel variants, and frequently fosters inflated claims regarding individual athletic prediction. Here, we present a population-calibrated, multi-tiered computational framework that evaluates genomic variation across six evidence streams integrating deep learning protein and genomic language models (ESM-2, Evo 2), missense and splice predictors (AlphaMissense, SpliceAI), and regional pathogenic-density metrics to generate a continuous Variant Priority Score (VPS; 1–10 scale). By dynamically renormalizing missing evidence streams rather than zero-filling them, the framework robustly scores unannotated variants lacking empirical database entries. Individual trait burden across 344 genes (spanning 17 physiological traits in Aerobic, Anaerobic, and Recovery domains) is aggregated and normalized against ancestry-matched expectations derived from gnomAD South Asian and an in-house reference cohort (N > 13,000). When applied to 960 South Indian individuals (584 male, 376 female; whole-exome sequencing), this normalization removed the dependence of expected trait burden on panel size by construction, and gene count explained only 2.7% of the variance in the resulting capacity scores (six traits interrogated by 16 to 33 genes). Variance partitioning of 4,764 individual-by-trait scores (six of the 17 traits: one aerobic, two anaerobic and three recovery) revealed that score variation was predominantly trait-driven (45.5%) rather than individual-driven (11.1%, p = 0.386), a pattern reproduced independently in males and females and with no statistically significant gender difference in capacity scores, indicating that, in this cohort, the candidate panel resolved differences between physiological traits but did not detect differences between individuals in capacity scores (no athletic phenotype was measured). Benchmarking against ClinVar classifications achieved an AUC of 0.74, which fell to 0.62 when archive-dependent streams were excluded, directly quantifying the role of shared data provenance. Replacing the in-house frequency reference with gnomAD South Asian shifted absolute scores (median fold change 0.73) but preserved trait ordering (Spearman 0.96), so scores are positions against a stated, population-specific reference. This framework provides an auditable, evidence-traceable baseline for trait-level research and functional variant prioritization while offering a critical methodological safeguard against over-interpreting unvalidated individual performance predictions.

## 1. INTRODUCTION

Human physiological response to standardized exercise protocols demonstrates profound inter-individual variability. In the landmark HERITAGE Family Study, 20 weeks of supervised endurance training yielded maximal oxygen uptake (VO_2_max) gains ranging from −114 to 1,097 mL/min **[3]**. Familial clustering of these response profiles confirmed a substantial hereditary contribution to trainability **[3]**. Subsequent investigations highlighted further biological complexity, demonstrating that exercise non-response is strongly dose-dependent **[5]** and that adverse metabolic adaptations occur in specific subgroups **[4]**.

Over two decades of candidate-gene association studies **[18]**, **[19]** have identified hundreds of physical performance markers **[7]**. Literature updates list up to 251 genetic markers reported to be associated with elite athletic status **[7]**, expanding from 155 identified in 2016 **[6]**. However, replication across independent cohorts remains exceptionally low: only 128 markers have been confirmed in at least two separate studies **[7]**, and systematic reviews of VO_2_max trainability loci show that, among the 97 genes identified, only 13 genetic variants have been reproduced by more than two research groups **[8]**. Meta-analyses demonstrate modest effect sizes for even the most extensively studied loci, such as *ACE* II genotype (OR = 1.23 for physical performance versus D-allele carriage) and *ACTN3* R577X RR genotype (OR = 1.21 in power disciplines) **[9]**. Large-scale international consortium studies comparing 1,520 endurance athletes against 2,760 controls failed to establish a shared polygenic profile **[10]**, underscoring the limited utility of simple candidate panels.

This lack of replication stems primarily from underpowered sample sizes and inconsistent phenotypic definition **[1]**, **[2]**. Whereas historical exercise genetics studies examined fewer than 1,000 subjects, contemporary genome-wide association studies (GWAS) of physical activity and grip strength pooling hundreds of thousands of participants show that complex physical traits are polygenic, governed by hundreds of loci with small individual effects **[13]**, **[14]**. While clear mechanistic links exist for individual variants such as *ACTN3* loss-of-function shifting muscle metabolism toward the aerobic pathway in mice **[11]** and the under-representation of the 577X allele in elite strength athletes **[12]** translating targeted panels into meaningful individual trait assessments presents major technical obstacles.

Conventional Direct-To-Consumer (DTC) reporting models **[20]** rely on Total Genotype Scores (TGS), an additive approach used in sport genomics **[21]**. TGS aggregates unweighted risk alleles across loci, introducing a critical design flaw: traits with higher panel gene counts artificially yield higher cumulative risk scores regardless of individual genetic architecture. Furthermore, lookup-dependent scoring skips unannotated novel variants, misinterpreting absence of empirical data as evidence of benign function. In this paper we evaluate a multi-tiered variant scoring framework against four specific claims, each tested and reported in §6: (1) variants absent from every annotation resource can still be scored from the evidence that exists; (2) normalizing against population-matched expectations reduces the dependence of a trait score on panel size; (3) the resulting capacity score resolves differences between traits and domains but not between individuals; and (4) the score ranks clinically pathogenic variants above benign ones, with a quantifiable share of that margin attributable to shared data provenance. Each is reported with its supporting evidence and with the limit of what that evidence establishes stated alongside it.

### 2. GENE LIST ARCHITECTURE & EVIDENCE TIERS

The evaluation framework encompasses 344 candidate genes mapped into 17 physiological traits spanning three key domains: Aerobic, Anaerobic, and Recovery (Table 1). Biological pathways were defined using GeneCards **[40]**, NCBI Gene **[43]**, Ensembl **[42]**, ClinVar **[23]**, UniProt **[41]**, PubMed, Reactome pathway annotations **[24]**. Gene-trait pairings were categorized into evidence confidence tiers (Top, Medium, Low) based on empirical literature support depth. Aerobic traits were deliberately divided into distinct sub-mechanisms (e.g., mitochondrial efficiency, capillary density) to preserve distinct physiological pathways.

**Table 1.** Panel Gene Distribution Across Traits and Evidence Tiers.

| **Domain** | **Physiological Trait** | **Genes** | **Top** | **Med** | **Low** |
|---|---|---|---|---|---|
| Aerobic | Mitochondrial Efficiency | 25 | 4 | 10 | 11 |
| Aerobic | Slow-Twitch Profile | 24 | 4 | 9 | 11 |
| Aerobic | Vascular / Capillary Density | 21 | 7 | 6 | 8 |
| Aerobic | Oxygen-Carrying Capacity | 18 | 3 | 9 | 6 |
| Aerobic | VO<sub>2</sub>max Potential | 9 | 4 | 4 | 1 |
| Aerobic | Gas Exchange / Lung Cap. | 6 | 2 | 3 | 1 |
| Aerobic | Cardiac-Output Efficiency | 2 | 0 | 2 | 0 |
| Anaerobic | Glycolysis | 23 | 8 | 10 | 5 |
| Anaerobic | Muscle Growth / Hypertrophy | 19 | 5 | 5 | 9 |
| Anaerobic | Lactate Buffering | 16 | 6 | 6 | 4 |
| Anaerobic | Fast-Twitch Profile | 11 | 4 | 4 | 3 |
| Anaerobic | ATP-PCr System | 6 | 4 | 1 | 1 |
| Recovery | Inflammatory Response | 43 | 9 | 16 | 18 |
| Recovery | Oxidative Stress Handling | 37 | 8 | 13 | 16 |
| Recovery | Bone Health / Osteoporosis | 33 | 8 | 13 | 12 |
| Recovery | Osteoarthritis Susceptibility | 28 | 6 | 7 | 15 |
| Recovery | Muscle Damage Risk | 23 | 8 | 7 | 8 |
| **Total** | **17 Traits (Unique: 344)** | **344** | **90** | **125** | **129** |

Evidence strength varies markedly across domains. Recovery traits feature the largest gene counts but possess the weakest experimental validation. A systematic review of competitive athletes identified only a small number of variants associated with soft-tissue injury susceptibility **[15]**, with exercise-induced muscle damage genetics remaining exploratory **[17]**. The single most robust association in this domain is *COL5A1* and anterior cruciate ligament rupture risk in females **[16]**.

## 3. VARIANT PRIORITY SCORING ENGINE

Variant functional effects are classified using the OpenCRAVAT annotation pipeline **[26]**. Each variant receives a Variant Priority Score (VPS; 1–10 scale) computed as a weighted average across six evidence streams (Table 2).

**Table 2.**
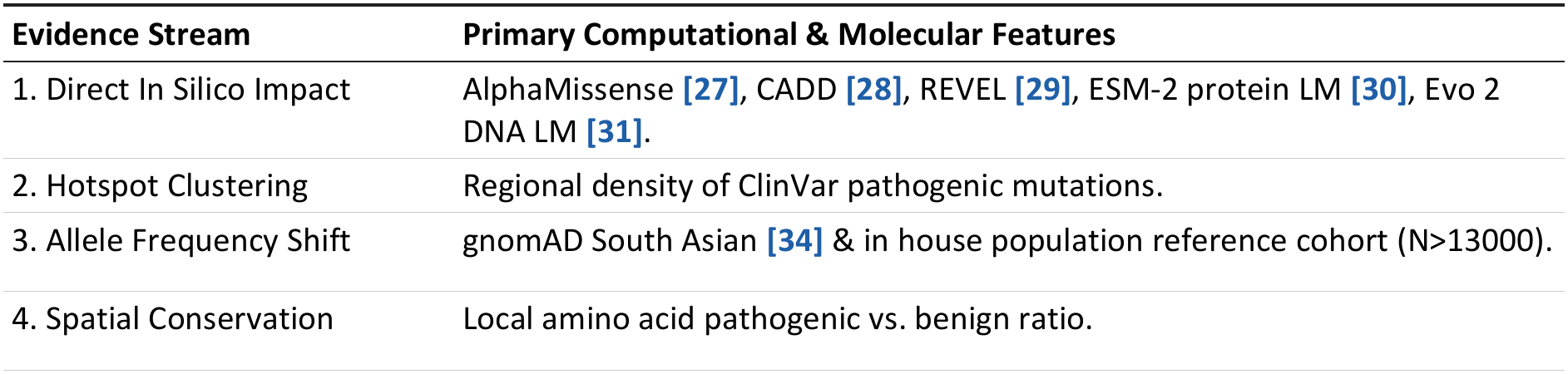
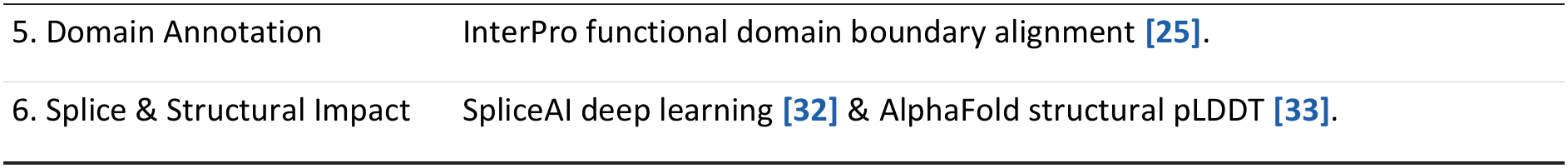
Evidence Streams for Single-Variant Priority Scoring.

Where a stream carries no data for a variant, its weight is redistributed pro rata over the populated streams and is never imputed. These weights are population- and panel-specific: they were calibrated for this 344-gene panel and for the South Asian frequency reference used by the allele-frequency stream, and they are not intended to be universal. Applying the framework to a cohort of different ancestry or to a different gene panel requires rebuilding the frequency reference and re-evaluating the weights (§5, §8). The six stream weights are: direct in silico impact 0.45, hotspot clustering 0.20, allele-frequency shift 0.15 (split equally 0.075/0.075 between the cohort and ancestry-matched sources), spatial conservation 0.10, domain annotation 0.05, splice and structural impact 0.05.

High-confidence SpliceAI splice disruption predictions **[32]** or expert-reviewed ClinVar pathogenic classifications **[23]** apply high-damage overrides. Population allele frequency filtering ensures common variants (AF > 0.05) are capped, following the ACMG/AMP stand-alone benign frequency criterion **[22]** and the maximum credible allele frequency framework **[35]**.

A novel missense variant in *BCL11A* (chr2) illustrates this dynamic handling: despite lacking population or clinical database entries, available in silico prediction tools, local hotspot proximity, and spatial conservation streams yielded a VPS of 6.65, demonstrating robust scoring without database dependency.

## 4. TRAIT NORMALIZATION & CAPACITY SCORING

The per-variant VPS (1–10) is converted to a damage coefficient *D* (0–1 scale). Individual trait damage is aggregated using zygosity multipliers (1 for heterozygous, 2 for homozygous alt) and gene-trait evidence weights. Trait damage is normalized against population expectations:

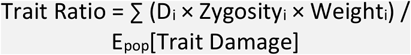

where *E*_*pop*_ represents expected damage calculated from an ancestry-matched reference population **[36]**. This ratio is mapped to a preserved capacity score (1–9 scale) by capacity = parity − min(1, log2(ratio)/log2(worst)) × (parity − floor) for ratio ≥ 1, and capacity = parity + min(1, log2(1/ratio)/log2(1/best)) × (ceiling − parity) for ratio < 1, with parity = 8.5, ceiling = 9.0, floor = 1.0, worst = 2048 and best = 0.5. Ratios below parity therefore grade continuously up toward the ceiling and ratios above parity grade continuously down toward the floor, so that no score is pinned at the ceiling, and matching population expectation yields an upper-middle baseline score (8.5 of 9.0) that avoids false ideal ceiling claims. All capacity figures in this manuscript use this mapping.

## STUDY COHORT, SEQUENCING & REPORTING STANDARDS

### Study population

The cohort comprised 960 individuals of South Indian ancestry who were undergoing preventive wellness screening as consumers. Participants were not selected on athletic status, training history or any performance phenotype, and none underwent a direct physiological assessment (§8). Demographic characteristics, including the count and percentage of males and females and the age distribution, are given in Table 3. Participants are identified by anonymised codes only; no recruitment-channel or site identifiers are reported.

**Table 3.** Characteristics of the Study Cohort (n = 960)

| **Characteristic** | **Pooled (n = 960)** | **Male (n = 584)** | **Female (n = 376)** |
|---|---|---|---|
| Individuals analysed, n (%) | 960 (100%) | 584 (60.8%) | 376 (39.2%) |
| Age, mean ± SD (years) | 46.7 ± 12.6 | 47.2 ± 12.4 | 45.9 ± 12.9 |
| Age, median (IQR) (years) | 46 (38–56) | 47 (39–56) | 45 (37–55) |
| Age, range (years) | 18–80 | 18–79 | 18–80 |
| Scoreable in aerobic domain, n (%) | 947 (98.6%) | 577 (98.8%) | 370 (98.4%) |
| Scoreable in anaerobic domain, n (%) | 959 (99.9%) | 584 (100.0%) | 375 (99.7%) |
| Scoreable in recovery domain, n (%) | 960 (100.0%) | 584 (100.0%) | 376 (100.0%) |
| Individual-by-trait capacity scores | 4,764 | 2,908 | 1,856 |
| Sequencing assay | WES | WES | WES |
| Reference genome | GRCh38 | GRCh38 | GRCh38 |

Percentages for gender are of the pooled cohort. Age is at sampling and is summarised for the 942 participants aged 18 years or older (572 male, 370 female, the male and female columns use the same 584/376 split throughout §6. Domain-level scoreable counts refer to individuals scoreable in that domain. The 4,764 individual-by-trait scores are the number of scoreable traits summed over individuals for the six traits analysed (§6), 4,734 across 955 individuals plus 30 from the remaining five individuals scored on all six, and are therefore not the product of the domain counts and the number of traits, because not every individual is scoreable for every trait.

### Sequencing and variant calling

All participants underwent whole-exome sequencing (WES). Sequencing was performed on an Illumina NovaSeq platform to an average on-target depth of 120× using the Twist exome capture panel, version 2.0. Reads were aligned to the GRCh38 reference genome with BWA-MEM **[38]** and variants were called with DeepVariant **[39]**. Functional consequences, population frequencies and in silico pathogenicity scores were annotated solely with an OpenCRAVAT-based pipeline **[26]** applied to an in-house cohort export. The South Asian frequencies used by the allele-frequency stream are from gnomAD v4.1.1 (GRCh38 exomes) **[34]**; no GRCh37 liftover was required. The present analysis is strictly downstream of this callset; full pipeline parameters and capture-kit quality-control metrics are held in the upstream cohort-generation protocol and are available from the corresponding author.

### Population specificity of the framework

Three components of the framework are specific to the South Asian setting in which it was built. First, the allele-frequency stream uses only ancestry-matched sources, gnomAD South Asian **[34]** and an in-house population reference (N > 13,000); all-ancestry gnomAD frequencies are not used in scoring. Second, the expected damage in the denominator of the trait ratio (§4) is computed from the same ancestry-matched frequencies, so a capacity score is a position relative to a South Asian expectation and not an absolute measure of function. Third, the stream weights (Table 2) were calibrated for this panel and this reference. Applying the framework to a cohort of different ancestry therefore requires rebuilding the frequency reference and re-examining the weights **[36]**. gnomAD South Asian pools several sub-populations, so frequencies in a South Indian cohort may differ from the pooled values; the in-house reference is intended to reduce this mismatch and cannot remove it.

## 6. RESULTS

The framework was applied to a cohort of 960 South Indian individuals, generating 4,764 individual-by-trait capacity scores (cohort characteristics in Table 3). Findings are reported below under four headings, corresponding to the four claims set out in §1. For each, we give the observation, the statistical evidence supporting it, and the boundary beyond which that evidence does not extend. The individual-level analyses in this section (domain distributions, variance partitioning and gender stratification) use the 4,764 individual-by-trait scores retained for six traits: Mitochondrial Efficiency (aerobic); Lactate Buffering and Muscle Growth (anaerobic); and Muscle Damage Risk, Bone Health and Osteoarthritis Susceptibility (recovery). The six traits comprise one aerobic, two anaerobic and three recovery traits, so the domain comparisons below compare one, two and three traits respectively, and the aerobic estimate rests on Mitochondrial Efficiency alone. The final subsection reports the comparison between males and females.

### Scoring of Unannotated and Novel Variants

The scoring engine returned a value for every variant on which at least one evidence stream carried data. Streams lacking data for a given variant were renormalized out of the weighted mean rather than assigned a value of zero, so that the denominator reflected only the evidence actually available. This treatment is deliberate: a zero would be indistinguishable from a prediction of tolerance, whereas the two statements no predictor evaluated this variant, and every predictor evaluated it as benign carry opposite implications. Variants on which no stream carried data received no score rather than a default one.

The practical consequence is illustrated by the BCL11A missense variant described in §3, which is recorded in no population database and carries no ClinVar classification **[23]**. A lookup-dependent pipeline returns nothing for such a variant and, in most reporting models, silently omits it. Here three streams were populated direct in silico impact **[27]**, **[28]**, **[30]**, hotspot clustering (nearest annotated pathogenic position 2 bp distant), and spatial conservation (no net enrichment) while the allele-frequency, domain-annotation and structural streams were renormalized out, yielding a VPS of 6.65. Had the three silent streams been zero-filled, the disruption signal carried by the populated streams would have been diluted and the variant would have been reported as substantially milder than the available evidence indicates.

Coverage of the allele-frequency stream was near-complete across the panel: approximately 0.2 - 0.3% of variants were absent from the in-house population reference cohort and were assigned the gnomAD South Asian frequency alone **[34]** rather than the two-source average. Across domains, per-variant scores clustered at the benign end of the 1-10 scale, with median values of 1.90 for both the aerobic (n = 401 variants, 105 genes) and anaerobic (n = 329, 75 genes) domains and 2.20 for recovery (n = 831, 164 genes); fewer than 4% of variants in any domain reached 5.0 (Fig. 1a-c). Variant counts are restricted to the 344 panel genes. The domains were statistically separable but practically similar (Kruskal-Wallis p = 0.0003), with recovery differing from aerobic (p = 0.0002) and anaerobic (p = 0.008) while aerobic and anaerobic did not differ from one another (p = 0.70). The recovery elevation of 0.30 median points tracks that domain’s higher proportion of protein-altering consequences rather than any property of the scoring rule.

**Fig. 1.**
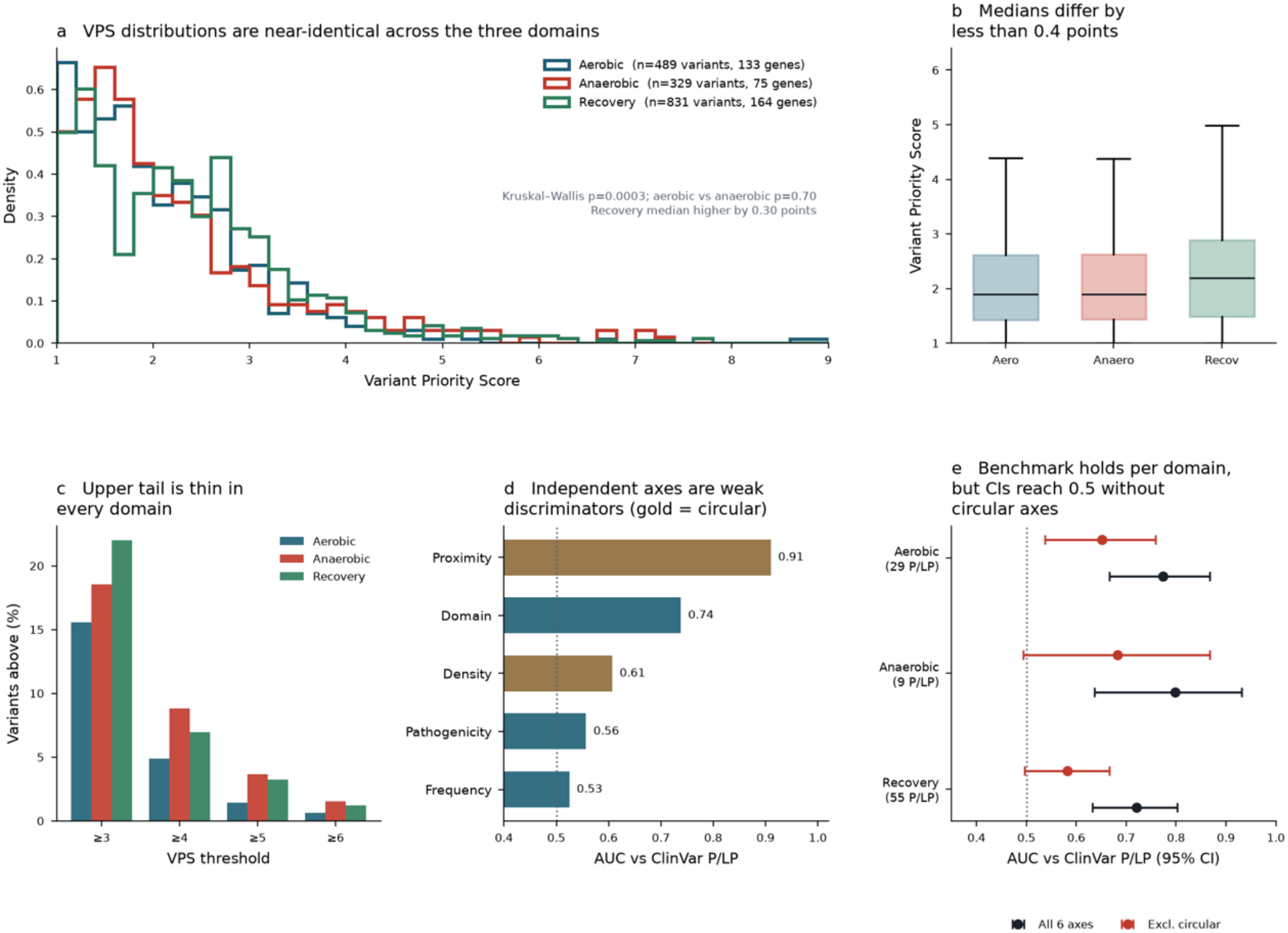
Per-domain VPS distributions and an external ClinVar benchmark. a, Density of per-variant VPS for aerobic (n = 401 variants, 105 genes), anaerobic (n = 329, 75 genes) and recovery (n = 831, 164 genes) domains; Kruskal–Wallis p = 0.0003, aerobic versus anaerobic p = 0.70. b, Domain medians (centre lines), interquartile ranges (boxes); medians differ by less than 0.4 points. c, Percentage of variants above VPS thresholds of 3, 4, 5 and 6; the upper tail is thin in every domain. d, AUC against ClinVar pathogenic/likely pathogenic versus benign/likely benign variants for five of the six single streams; gold bars mark streams computed from the ClinVar archive that also supplies the labels; dotted line, chance (0.5). e, AUC with 95% bootstrap confidence intervals (2,000 resamples) per domain for all six streams (black) and for the archive-independent streams only (red); numbers of pathogenic/likely pathogenic variants are shown in brackets.

This result establishes coverage, not accuracy. Because silent streams are renormalized rather than imputed, two variants may receive identical scores while resting on different numbers of populated streams, so scores are not of uniform precision and the supporting stream count should be reported alongside the value.

### Dependence of Trait Scores on Panel Composition

An unweighted allele sum rises with the number of variants a panel interrogates for a given trait, so a cross-trait comparison within one individual partly measures the panel’s own composition rather than that individual’s genotype **[21]**. Dividing the observed weighted damage by the damage expected from ancestry-matched allele frequencies **[34]**, **[36]** removes panel content from numerator and denominator alike, since both are computed over the same variant set. In the present panel the contrast is large: Inflammatory Response is interrogated by 43 genes and Cardiac-Output Efficiency by 2 (Table 1), a more than twenty-fold difference in the number of genes that can contribute damage to the trait.

The normalization removes this dependence by construction: the expected burden in the denominator grows with the same variant set as the observed burden in the numerator, so the scale of the ratio does not depend on panel size (this is a statement of scale invariance and not of the cohort median, which need not equal one; see Population-Specific Calibration), whereas the expectation of an unweighted sum grows in proportion to it. Empirically, in the analysis table retained for this cohort (six traits, each interrogated by 16 to 33 genes), trait gene count explained 2.7% of the variance in individual-by-trait capacity scores after normalization (Pearson r = 0.16, n = 4,764 scores) and was not correlated with trait mean score (Spearman ρ = 0.14, p = 0.79, n = 6 traits). This check spans 16 to 33 genes and does not include the extremes cited above (Cardiac-Output Efficiency, 2 genes; Inflammatory Response, 43 genes), so it does not test the full range of panel sizes.

A residual structure survives normalization and constrains how the result may be read. Within an individual, the rank order of trait scores correlated positively with the median allele frequency of each trait’s variants, indicating that cross-trait ordering continues to track the frequency structure of panel content rather than physiology. Normalization therefore supports a statement about one trait in isolation; it does not license the ranking of one individual’s traits against one another.

### Distribution of Capacity Scores and Partitioning of Variance

Two analyses bear on whether the capacity score distinguishes individuals, and they converge on the same answer. Partitioning the variance of the 4,764 individual-by-trait scores attributed 45.5% to trait identity and 11.1% to individual identity, with the individual term not reaching statistical significance (p = 0.386; two-way additive model, capacity ∼ trait + individual, Type II sums of squares). An individual share of this size is what residual variation alone would produce (F ≈ 1.0 on 959 and 3,799 degrees of freedom), so it is read as no detectable individual-level signal. Independently, the capacity distributions of the three domains separated clearly from one another, although by small absolute margins (domain medians 8.12, 7.66 and 7.85; Kruskal-Wallis p = 1.8 × 10^−92^, all pairwise contrasts p < 10^−8^; Table 4; Fig. 2). Because the six traits comprise one aerobic, two anaerobic and three recovery traits, this domain contrast compares one, two and three traits. This decomposition was repeated independently within the male and female subsets (§6, Gender-Stratified Analysis) and reproduces the same pattern in both.

**Table 4.** Preserved Capacity Score Distributions by Domain (1–9 Scale)

| **Domain** | **n (M)** | **Median (M)** | **Mean (SD) (M)** | **n (F)** | **Median (F)** | **Mean (SD) (F)** | **Mann–Whitney p (M vs F)** |
|---|---|---|---|---|---|---|---|
| Aerobic | 577 | 8.12 | 8.15 (0.42) | 370 | 8.10 | 8.15 (0.37) | 0.63 |
| Anaerobic | 584 | 7.63 | 7.31 (1.23) | 375 | 7.70 | 7.35 (1.15) | 0.93 |
| Recovery | 584 | 7.85 | 7.77 (0.45) | 376 | 7.85 | 7.74 (0.50) | 0.60 |

**Fig. 2.**
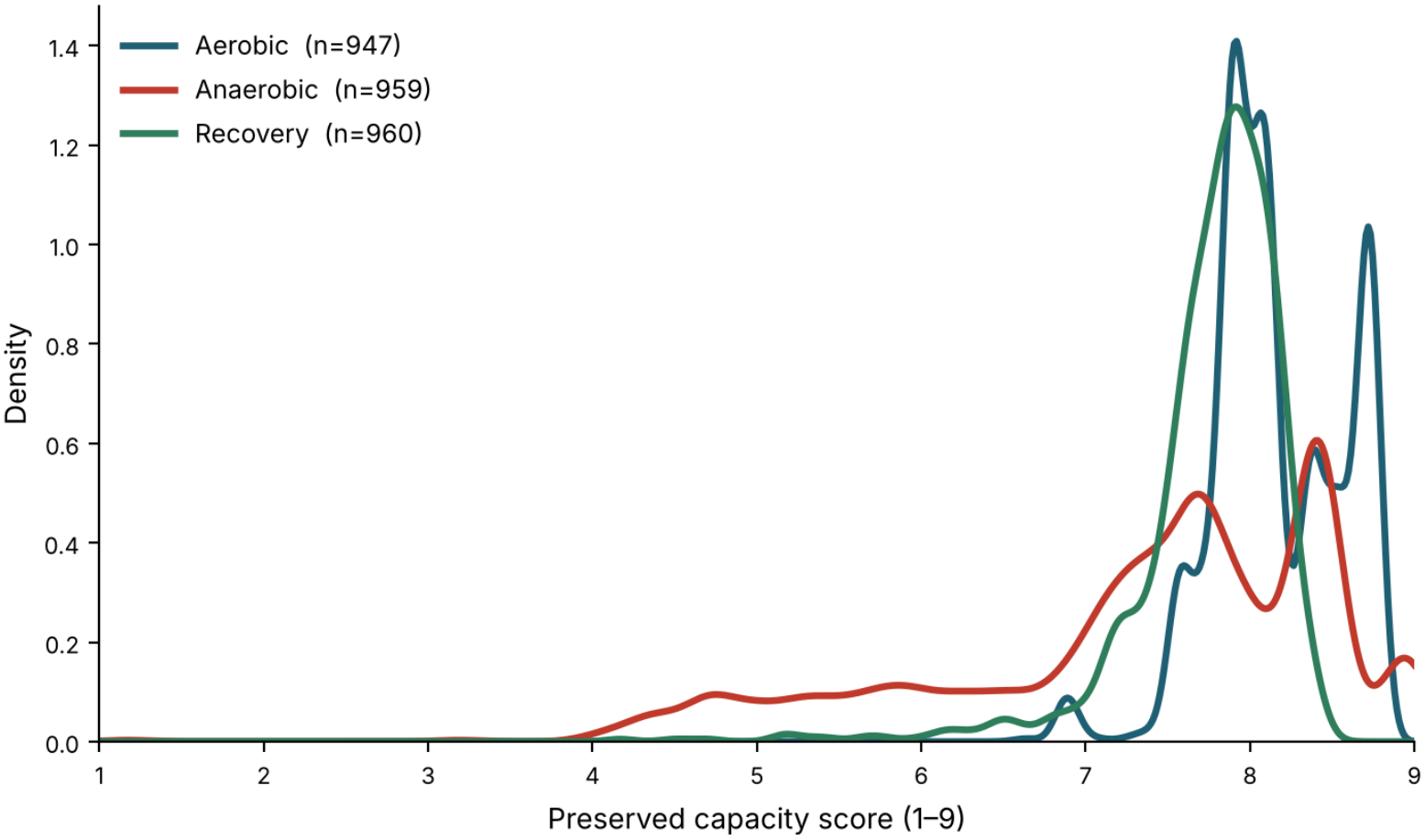
Distribution of preserved capacity scores by domain. Kernel density estimates for aerobic (n = 947), anaerobic (n = 959) and recovery (n = 960) domains, under the anchored mapping (parity = 8.5, ceiling = 9.0, floor = 1.0; §4). The x axis shows the capacity score on its native 1-9 scale; Table 4 reports the same scale. No domain shows ceiling pinning under this mapping (maximum observed scores 8.77 aerobic, 8.94 anaerobic, 8.42 recovery).

These findings are frequently misread as contradictory and are not. Both locate the resolvable structure on the same axis. A capacity score shifts substantially when the trait or domain under examination changes, and shifts little when the individual changes. The framework therefore resolves the axis it was constructed to resolve which physiological mechanism is being interrogated and does not resolve the axis a consumer-facing report would require, namely which individual is being examined.

The distributions depart sharply from uniformity (Kolmogorov–Smirnov against a uniform distribution on the reported range, p < 10^−40^ for each), which follows from the construction rather than indicating an artefact: parity with the population expectation is anchored toward the upper end of the scale (8.5 of 9.0), so a cohort drawn from that same population necessarily occupies the upper half. A reported value is therefore a position relative to a population expectation and not a percentile rank within this cohort. Under the anchored mapping no domain shows ceiling pinning (maximum scores 8.77 aerobic, 8.94 anaerobic, 8.42 recovery; §4). As a robustness check, the two deliberately disjoint effect-weight schemes described in §4 one anchored on clinical classification **[23]**, the other using computational predictors exclusively were carried through the full pipeline and agreed closely, indicating that the partitioning result does not depend on which weighting was adopted. Capacity scores did not differ between males and females in any domain (Table 4; Mann–Whitney p = 0.63 aerobic, p = 0.93 anaerobic, p = 0.60 recovery).

What follows from this is a restriction rather than a capability. The framework does not support ranking an individual’s traits against one another, which the residual frequency structure described above independently precludes, nor comparing individuals with one another, which a non-significant individual term precludes directly.

### Population-Specific Calibration

Capacity scores in Table 4 are positions relative to the South Asian expectation defined in §4 and §5, and should be read as such. To quantify how far the result depends on that choice, the expectation denominator was recomputed against an alternative frequency reference while every other quantity was held fixed: the in-house South Indian cohort frequency was replaced by the gnomAD South Asian frequency for the same variants. The analysis covers 16,221 individual-by-trait observations from 958 individuals across all 17 traits. Substituting the reference lowered the median observed-to-expected ratio from 0.324 to 0.237, a median fold change of 0.73, because the gnomAD South Asian panel carries systematically higher frequencies for these variants than the in-house cohort and therefore raises the expected burden. The effect is close to uniform in rank: the two sets of ratios correlate at Spearman 0.961 across all observations, and within an individual the rank order of traits is preserved at a median Spearman of 0.912 (interquartile range 0.870-0.943), with the single highest-ratio trait unchanged in 667 of 958 individuals (69.6%). The shift is not uniform across traits: it is largest for Oxygen-Carrying Capacity (fold change 0.57) and smallest for the ATP-PCr system (0.89), so the absolute capacity value attached to a trait is reference-dependent even though the ordering is largely not. The practical reading is that these scores are positions against a stated reference and that cross-study comparison requires the reference to be stated with them; they are not portable constants.

### Agreement With Clinical Pathogenicity Classification

A reference set was assembled independently of the cohort from all classified variants in the panel’s genes **[23]**, comprising 25,404 Pathogenic or Likely Pathogenic and 69,259 Benign or Likely Benign entries. Evaluation was performed on the 503 cohort variants matching a classified position (93 pathogenic, 410 benign). The full six-stream score achieved an area under the receiver-operating characteristic curve of 0.74, with mean scores of 2.88 for pathogenic against 2.09 for benign variants. The ranking is therefore in the expected direction, and the substantive question is how much of that separation rests on evidence independent of the labels being predicted.

Decomposing the score by stream answers this directly. Hotspot clustering, which is computed from the positions of archive-annotated pathogenic variants, was the strongest single discriminator at 0.91 (Fig. 1d). Domain annotation **[25]** reached 0.74 and is independent of the benchmark. Spatial conservation, also archive-derived, reached 0.61. The direct in silico impact stream the only one that evaluates the substitution itself, and the most heavily weighted of the six reached 0.56, and allele frequency reached 0.52. The strongest-performing stream is therefore one whose provenance is shared with the answer key, and the second-strongest, domain annotation, is independent of it, while the stream carrying the greatest weight in the composite score performs barely above chance against this particular reference. Recomputing the score over only the streams that draw on no clinical archive, renormalized as in §3, gives 0.62 against 0.74 for the full score, so approximately half of the margin above chance is independent evidence and approximately half is shared provenance. The ClinVar-pathogenic high-damage override described in §3 was active when these AUCs were computed: for variants whose ClinVar review status reaches the definitive tier, the ClinVar-derived weight replaces the predictor-based weight. Hotspot clustering and spatial conservation are not computed leave-one-out: both count every matching reference position without excluding the query variant’s own record, so a benchmark variant that is itself present in the reference archive contributes to its own proximity and density scores.

The same decomposition repeats within every domain (Table 5; Fig. 1e). Each exceeds chance on the full score and each loses a comparable amount when the archive-derived streams are withdrawn, establishing the effect as a property of how the composite score is constructed rather than of any single domain’s gene content. On independent evidence alone, only the aerobic domain clearly excludes chance; the anaerobic estimate rests on nine classified pathogenic variants and its confidence interval spans 0.5.

**Table 5.**
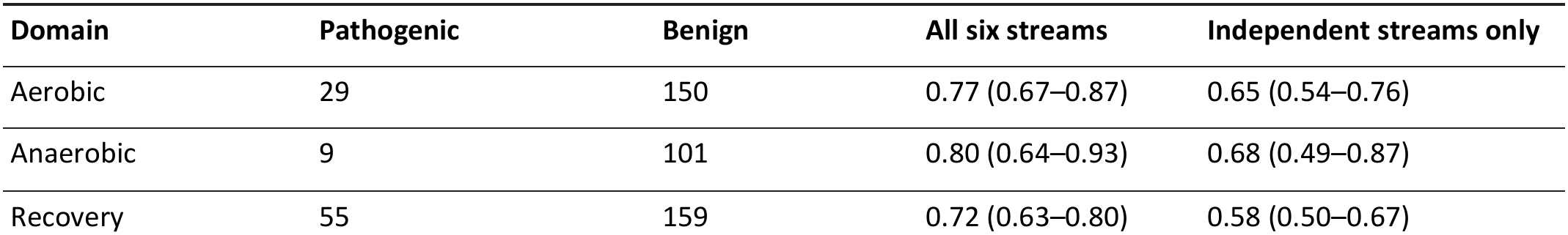
Discrimination Against Clinical Classification by Domain, Full Score Versus Archive-Independent Streams (95% confidence intervals from 2,000 bootstrap resamples)

| Domain | Pathogenic | Benign | All six streams | Independent streams only |
|---|---|---|---|---|
| Aerobic | 29 | 150 | 0.77 (0.67–0.87) | 0.65 (0.54–0.76) |
| Anaerobic | 9 | 101 | 0.80 (0.64–0.93) | 0.68 (0.49–0.87) |
| Recovery | 55 | 159 | 0.72 (0.63–0.80) | 0.58 (0.50–0.67) |

This agreement does not validate the capacity score. A ClinVar classification records pathogenicity for Mendelian disease under ACMG/AMP criteria **[22]**, whereas the capacity score describes preserved physiological function; a variant may be benign for monogenic disease while still shifting a quantitative physiological trait, and the converse holds equally. The benchmark establishes that the pathogenicity-adjacent streams behave coherently against an external reference, and establishes nothing further.

### Gender-Stratified Analysis

The analyses were repeated separately in the 584 male and 376 female participants (gender as recorded in the cohort metadata; Table 3). The two groups were similar in age (median 47 versus 45 years, p = 0.09) and in the share of individuals scoreable in each domain (within 0.4 percentage points).

Capacity scores did not differ between males and females in any domain (aerobic p = 0.63, anaerobic p = 0.93, recovery p = 0.60; Table 6 and Fig. 3b). For the six individual traits, no difference remained after correction for multiple testing (smallest corrected p = 0.22, Lactate Buffering).

**Table 6.** Capacity Score by Gender and Domain (1–9 scale)

| Domain | Male, median (IQR; n = 577–584) | Female, median (IQR; n = 370–376) | Mann–Whitney p |
|---|---|---|---|
| Aerobic | 8.12 (7.89–8.54) | 8.10 (7.89–8.46) | 0.63 |
| Anaerobic | 7.63 (6.74–8.42) | 7.70 (7.02–8.32) | 0.93 |
| Recovery | 7.85 (7.60–8.06) | 7.85 (7.63–8.02) | 0.60 |

**Fig. 3.**
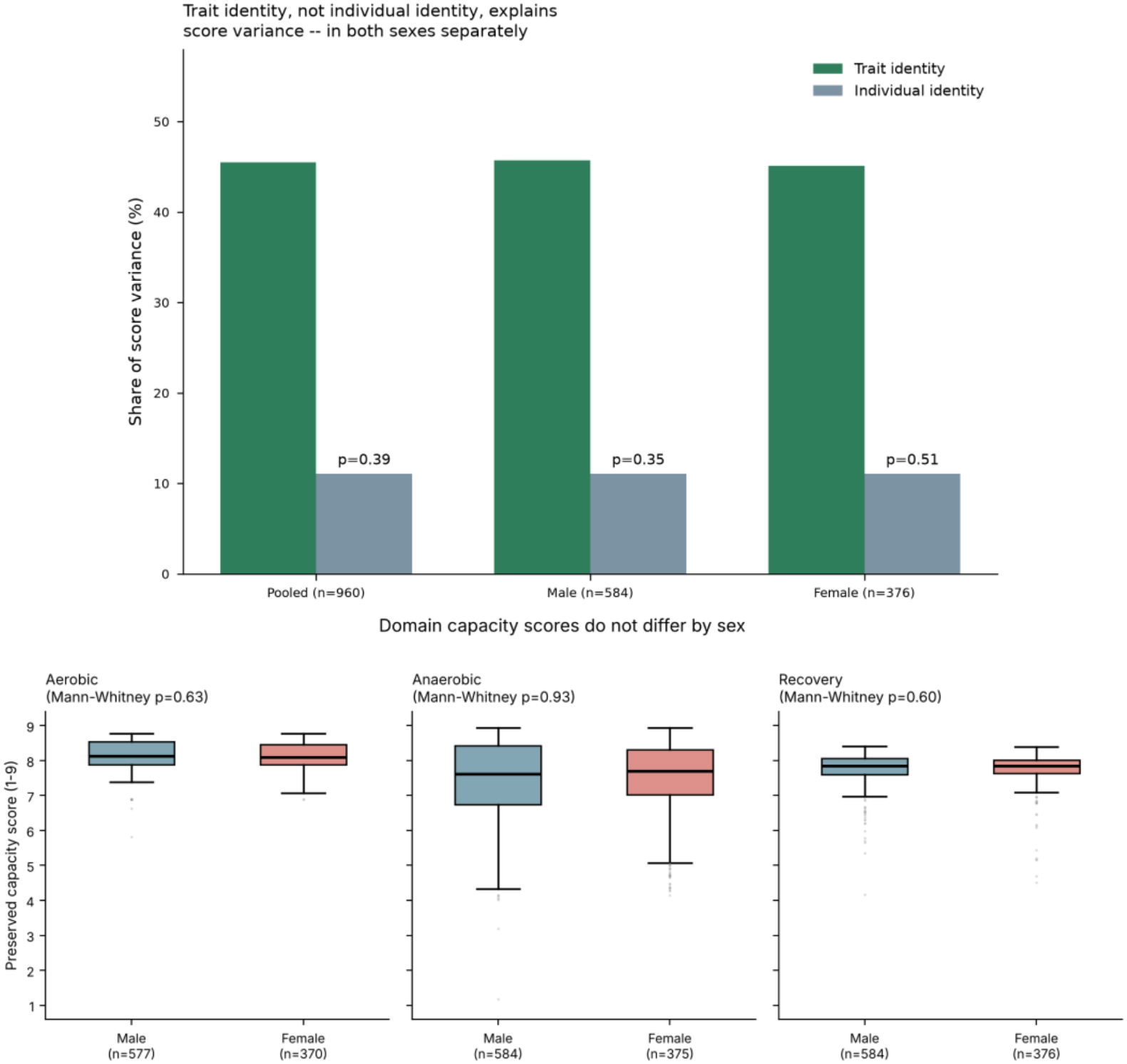
Gender-stratified results. a, Share of score variance attributed to trait identity versus individual identity, computed independently for the pooled cohort, males only, and females only. b, Preserved capacity-score distributions by domain, split by gender, with the p-value for the male-versus-female comparison in each domain.

The main finding also held within each gender: trait identity explained 45.7% of score variance in males and 45.1% in females (pooled 45.5%), and individual identity explained 11.1% in both, not significant in either (Fig. 3a).

Table 6 gives the domain medians for each gender.

In this cohort, therefore, gender did not have a statistically significant effect on capacity scores: the framework gave the same answer in men and women, and we found no evidence of gender bias in the scoring. The female group (n = 376) is smaller than the male group, so small differences cannot be excluded.

Twelve of the 344 panel genes are X-linked. Compared gene by gene across all 344 genes, carrier rates differed between males and females for no autosomal gene after correction; the two genes that did differ, TIMP1 and FOXP3, are X-linked and reflect the genotype-calling limitation described in §8 rather than biology.

## 7. DISCUSSION

We developed a population-referenced, evidence-weighted framework that converts a curated gene panel into trait-level capacity scores, and applied it to 960 South Indian individuals. The findings differ in strength, and we discuss each at the level the evidence supports. In brief, the framework scores variants that lookup-based pipelines cannot, it reduces the influence of panel composition, it resolves differences between physiological traits and domains, and it ranks clinically classified variants in the expected direction. We interpret these results as properties of a research instrument for trait-level analysis and not as a predictor of individual performance.

### Resolution at the level of traits and domains

Trait identity accounted for 45.5% of score variance, and the three domains separated clearly (Kruskal–Wallis p = 1.8 × 10^−92^), although by small absolute margins (domain medians 8.12, 7.66 and 7.85). The capacity score therefore captures which physiological mechanism is under examination, and Table 4 shows that the domains differ in centre and spread, the anaerobic domain being the widest. This is the axis the framework was designed to resolve, and it is the one that supports pathway-level interpretation, such as identifying which mechanisms carry the greatest variant burden in a cohort or sub-cohort. The individual term was small (11.1%) and not statistically significant (p = 0.386), so individuals in this cohort were similar to one another. That is the expected pattern for a panel of 344 genes whose members each have small effects. Large GWAS of physical activity and grip strength find that these traits are governed by many loci of small individual effect **[13]**, **[14]**, and replication of candidate markers for athletic status and VO2max trainability remains limited **[6]**–**[8]**, **[10]**. A score that separated individuals sharply from such a panel would be at odds with that evidence. The agreement between the two weighting schemes, one anchored on clinical classification and one using computational predictors only, indicates that this structure is not an artefact of a particular weighting choice. This finding did not depend on gender: the same decomposition gave the same answer within males and within females, and capacity scores did not differ significantly between them in any domain (§6, Gender-Stratified Analysis).

### Normalization and the scoring of novel variants

Dividing observed damage by ancestry-matched expected damage removes panel content from both numerator and denominator and reduced the association between a trait’s score and the number of variants assayed, which is the main weakness of total genotype scores **[21]**. In the retained analysis table, trait gene count explained only 2.7% of capacity-score variance after normalization in six traits of 16 to 33 genes (§6). The renormalization rule gave the framework a second practical advantage. A variant absent from every population and clinical resource, such as the BCL11A missense variant, still received a score (VPS 6.65) from the streams that carried data, and silent streams were removed from the weighted mean rather than zero-filled. This keeps the absence of evaluation separate from evidence of tolerance, which is the point at which lookup-dependent pipelines most often fail for poorly characterized variants. The price is that precision is not uniform across variants, so we recommend reporting the number of contributing streams alongside each score.

### Agreement with clinical classification

The full score reached an AUC of 0.74 against ClinVar, and 0.62 when computed only from streams independent of the archive. The score therefore ranks pathogenic variants above benign ones using evidence beyond the answer key, and the gain from archive-derived streams is itself informative. The stream-level decomposition shows that hotspot clustering was the strongest discriminator (0.91), while the direct in silico impact stream, which carries the most weight in the composite, reached 0.56. This identifies a concrete route to improvement: reweighting or recalibrating the direct-impact stream, and testing the label-independent streams against deep mutational scanning data where it exists **[37]**. The decomposition was consistent across all three domains (Table 5). We note that ClinVar records Mendelian pathogenicity under ACMG/AMP criteria **[22]**, whereas the capacity score describes quantitative physiological function, so the benchmark tests the coherence of the pathogenicity-adjacent streams and not the capacity score itself. A modest AUC against a reference built for a different question is to be expected.

### Provenance and auditability

The main contribution of this framework is specified provenance and not superior prediction. Each weighting layer acts at a declared stage, outputs are emitted before and after each layer, and every reported value traces to the evidence that produced it. Zygosity is kept out of the per-variant score and enters only as allele dosage at the burden stage, so that each variant is scored once and reused across all carriers. This allows a reader to isolate and test each tuned component, which direct-to-consumer performance products have been criticised for not permitting **[20]**.

### Gender

Capacity scores, variance partitioning and per-trait comparisons showed no statistically significant difference between males and females, so we found no evidence of gender bias in the scoring within this cohort. The female group is smaller (n = 376), and the 12 X-linked genes were genotyped without ploidy-aware correction (§8), so this is a statement about this cohort and callset and not a claim that gender is irrelevant to fitness genetics.

### Population specificity

Substituting the gnomAD South Asian frequencies for the in-house reference lowered the median observed-to-expected ratio from 0.324 to 0.237 (median fold change 0.73) but left the ordering largely intact (Spearman 0.961 across observations; median 0.912 within individuals; the top-ranked trait unchanged in 69.6% of individuals). Absolute capacity values are therefore positions against a stated South Asian reference, not population-free quantities, and the stream weights and frequency reference must be rebuilt before the framework is applied to another ancestry or panel.

### Intended use and governance

On this basis the framework is suited to trait-level burden profiling of cohorts, to prioritizing variants for functional follow-up, and to generating hypotheses about which pathways merit phenotypic testing. It should not be used to rank individuals or to predict fitness, trainability or athletic potential. The population-anchored distributions reported here (Table 4) also provide a reference for South Asian cohorts that is scarce in the literature, against which future sub-cohorts, such as trained and untrained groups, can be compared. Pathogenic and Likely Pathogenic findings in cardiomyopathy, myopathy and connective-tissue genes are routed to clinical review and not into the fitness readout **[22]**, **[23]**, and the possibility of such findings must be disclosed at consent. Together with the re-identifiability of genomic data and the weaker protection of raw data held by third-party services, this places the framework within the expectations of the FIMS consensus statements **[2]**, **[20]**.

### Future directions

The most informative next step is phenotypic: measuring VO2max or lactate threshold in a subset of this cohort would test whether trait-level scores track physiology and would supply per-trait reference distributions. Lower-cost steps include correlation against deep mutational scanning data **[37]**, systematic evaluation of the label-independent streams across the full panel, recomputation of the panel-size coefficient, and calibration against an ancestry-matched reference held out of score construction **[36]**. Until such validation exists, the framework should be regarded as a reproducible, evidence-traceable tool for research and prioritization.

## 8. LIMITATIONS

### No measured phenotype

No individual in this cohort underwent VO2max, lactate-threshold or any other direct physiological assessment. The capacity score has therefore never been tested against the quantity it purports to describe. This is the binding limitation, and no additional computational benchmarking substitutes for it **[1]**, **[8]**.

### Benchmark provenance is partly shared

Hotspot clustering and spatial conservation are computed from the same archive supplying the benchmark labels **[23]**, raising the measured AUC from 0.62 to 0.74. The per-domain decomposition shows this is structural rather than domain-specific, but it cannot be removed without an independent functional reference. The ClinVar high-damage override described in §3 is a third channel through which ClinVar enters the composite score and was active for the reported AUCs, so the archive-independent figure of 0.62 may itself be partly influenced by ClinVar. Hotspot clustering and spatial conservation are also not computed leave-one-out.

### Trait coverage

Individual-level analyses use six of the 17 traits (one aerobic, two anaerobic and three recovery). Domain contrasts are therefore contrasts of one, two and three traits, and the aerobic estimate rests on a single trait.

## 9. CONCLUSION

In complex trait genomics, the computational architecture that transforms variant calls into individual interpretations demands the same rigorous validation as panel gene selection. We have formulated a transparent, population-referenced framework that systematically evaluates candidate variants across six computational evidence streams, accommodates novel variants via dynamic denominator renormalization, and mitigates the gene-count artifacts inherent to conventional allele-counting methods. When evaluated across 960 South Indian individuals, the framework yielded five central findings:

### Robust novel variant scoring

Renormalizing unpopulated streams enables scoring of ultra-rare or novel variants without conflating an absence of annotation with predicted tolerance.

### Trait-level, not individual-level, discrimination

Score variance is overwhelmingly governed by physiological trait identity (45.5%) rather than individual genetic identity (11.1%, p = 0.386). The framework reliably characterizes mechanistic pathway burden across cohorts, but does not support ranking individuals against one another or predicting personal athletic performance.

### Quantifiable provenance inflation

While the model achieves an AUC of 0.74 against ClinVar pathogenic variants, half of its discriminative margin above chance (0.12 of 0.24) stems from shared provenance with clinical archives, underscoring the necessity of archive-independent validation.

### No gender bias in this cohort

Capacity scores, variance partitioning and per-trait comparisons did not differ significantly between males and females (all domain p ≥ 0.60), so we found no statistically significant gender bias.

### Population-specific calibration

Replacing the in-house frequency reference with gnomAD South Asian shifted absolute scores (median fold change 0.73) but preserved trait ordering (Spearman 0.96), so scores are positions against a stated reference and the framework must be recalibrated for other ancestries or panels.

Ultimately, computational scoring cannot substitute for empirical human physiology. The decisive validation of this framework will require linking trait capacity scores directly to measured physiological phenotypes, such as V?O_2_max trainability and blood lactate kinetics, alongside functional deep mutational scanning assays. Until such empirical validation is achieved, this framework should be utilized strictly as an auditable, hypothesis-generating tool for cohort-level pathway prioritization, establishing an ethical and reproducible alternative to uncalibrated direct-to-consumer performance metrics.

## Supporting information

PER PATIENT VPS SCORES

## Data Availability

De-identified score matrices and implementation scripts are available from the corresponding author upon reasonable academic request. Individual-level exome data are not publicly available because of participant privacy and consent restrictions.

## 10. DATA AVAILABILITY

De-identified score matrices and implementation scripts are available from the corresponding author upon reasonable academic request.

## 11. ETHICS

Participants were adults who underwent preventive wellness screening as consumers and informed consent was obtained from all participants; the analysis used de-identified exome data and anonymised participant codes.

## 12. AUTHOR CONTRIBUTIONS

Kalyan Ram Uppaluri and Krishna Kumar Vempati led the project. Hima Jyothi Challa guided the selection of the gene panel and the assignment of evidence tiers related to the genomic concepts. Jayanthy Prahashit extracted the candidate genes from the different source databases and websites. Balaji Malluri and Madhu Bhanu Varma Rayala annotated the data. Lakshmi Nirmala Kadali implemented the scoring algorithm and statistical analysis. All authors reviewed and approved the final manuscript.

## REFERENCES

1. Hagberg, J. M. et al. Advances in exercise, fitness, and performance genomics in 2010. Med. Sci. Sports Exerc. 43, 743–752 (2011).

2. Tanisawa, K. et al. Sport and exercise genomics: the FIMS 2019 consensus statement update. Br. J. Sports Med. 54, 969–975 (2020).

3. Bouchard, C. et al. Familial aggregation of VO2max response to exercise training: results from the HERITAGE Family Study. J. Appl. Physiol. 87, 1003–1008 (1999).

4. Bouchard, C. et al. Adverse metabolic response to regular exercise: is it a rare or common occurrence? PLoS ONE 7, e37887 (2012).

5. Montero, D. & Lundby, C. Refuting the myth of non-response to exercise training. J. Physiol. 595, 3377–3387 (2017).

6. Ahmetov, I. I. et al. Genes and Athletic Performance: An Update. Med. Sport Sci. 61, 41–54 (2016).

7. Semenova, E. A. et al. Genes and Athletic Performance: The 2023 Update. Genes 14, 1235 (2023).

8. Williams, C. J. et al. Genes to predict VO2max trainability: a systematic review. BMC Genomics 18, 831 (2017).

9. Ma, F. et al. The Association of Sport Performance with ACE and ACTN3 Genetic Polymorphisms. PLoS ONE 8, e54685 (2013).

10. Rankinen, T. et al. No Evidence of a Common DNA Variant Profile Specific to World Class Endurance Athletes. PLoS ONE 11, e0147330 (2016).

11. MacArthur, D. G. et al. Loss of ACTN3 gene function alters mouse muscle metabolism and shows evidence of positive selection in humans. Nat. Genet. 39, 1261–1265 (2007).

12. Roth, S. M. et al. The ACTN3 R577X nonsense allele is under-represented in elite-level strength athletes. Eur. J. Hum. Genet. 16, 391–394 (2008).

13. Wang, Z. et al. Genome-wide association analyses of physical activity and sedentary behavior provide insights into underlying mechanisms and roles in disease prevention. Nat. Genet. 54, 1332–1344 (2022).

14. Willems, S. M. et al. Large-scale GWAS identifies multiple loci for hand grip strength providing biological insights into muscular fitness. Nat. Commun. 8, 16015 (2017).

15. Appel, M. et al. Effects of Genetic Variation on Endurance Performance, Muscle Strength, and Injury Susceptibility in Sports: A Systematic Review. Front. Physiol. 12, 694411 (2021).

16. Posthumus, M. et al. The COL5A1 Gene Is Associated With Increased Risk of ACL Ruptures. Am. J. Sports Med. 37, 2234–2240 (2009).

17. Baumert, P. et al. Genetic variation and exercise-induced muscle damage. Eur. J. Appl. Physiol. 116, 1595–1625 (2016).

18. Bouchard, C. et al. Genetics of Fitness and Physical Performance (Human Kinetics, 1997).

19. Bouchard, C. & Hoffman, E. P. Genetic and Molecular Aspects of Sport Performance (Wiley-Blackwell, 2011).

20. Webborn, N. et al. Direct-to-consumer genetic testing for predicting sports performance and talent identification: Consensus statement. Br. J. Sports Med. 49, 1486–1491 (2015).

21. Pranckevičienė, E. et al. Total Genotype Score Modelling of Polygenic Endurance-Power Profiles in Lithuanian Elite Athletes. Genes 12, 1067 (2021).

22. Richards, S. et al. Standards and guidelines for the interpretation of sequence variants. Genet. Med. 17, 405–424 (2015).

23. Landrum, M. et al. ClinVar: improving access to variant interpretations and supporting evidence. Nucleic Acids Res. 46, D1062–D1067 (2018).

24. Fabregat, A. et al. The Reactome Pathway Knowledgebase. Nucleic Acids Res. 46, D649–D655 (2018).

25. Paysan-Lafosse, T. et al. InterPro in 2022. Nucleic Acids Res. 51, D418–D427 (2023).

26. Pagel, K. A. et al. Integrated informatics analysis of cancer-related variants. JCO Clin. Cancer Inform. 4, 310–317 (2020).

27. Cheng, J. et al. Accurate proteome-wide missense variant effect prediction with AlphaMissense. Science 381, eadg7492 (2023).

28. Rentzsch, P. et al. CADD: predicting deleteriousness of variants. Nucleic Acids Res. 47, D886–D894 (2019).

29. Ioannidis, N. M. et al. REVEL: An Ensemble Method for Predicting the Pathogenicity of Rare Missense Variants. Am. J. Hum. Genet. 99, 877–885 (2016).

30. Lin, Z. et al. Evolutionary-scale prediction of atomic-level protein structure with a language model. Science 379, 1123–1130 (2023).

31. Brixi, G. et al. Genome modelling and design across all domains of life with Evo 2. Nature 652, 1349–1361 (2026).

32. Jaganathan, K. et al. Predicting Splicing from Primary Sequence with Deep Learning. Cell 176, 535–548 (2019).

33. Jumper, J. et al. Highly accurate protein structure prediction with AlphaFold. Nature 596, 583–589 (2021).

34. Chen, S. et al. A genomic mutational constraint map using variation in 76,156 human genomes. Nature 625, 92–100 (2024).

35. Whiffin, N. et al. Using high-resolution variant frequencies to empower clinical genome interpretation. Genet. Med. 19, 1151–1158 (2017).

36. Martin, A. R. et al. Human Demographic History Impacts Genetic Risk Prediction across Diverse Populations. Am. J. Hum. Genet. 100, 635–649 (2017).

37. Fowler, D. M. & Fields, S. Deep mutational scanning: a new style of protein science. Nat. Methods 11, 801–807 (2014).

38. Li, H. Aligning sequence reads, clone sequences and assembly contigs with BWA-MEM. Preprint at arXiv https://arxiv.org/abs/1303.3997 (2013).

39. Poplin, R. et al. A universal SNP and small-indel variant caller using deep neural networks. Nat. Biotechnol. 36, 983–987 (2018).

40. Stelzer, G. et al. The GeneCards Suite: From Gene Data Mining to Disease Genome Sequence Analyses. Curr. Protoc. Bioinformatics 54, 1.30.1–1.30.33 (2016).

41. The UniProt Consortium. UniProt: the Universal Protein Knowledgebase in 2023. Nucleic Acids Res. 51, D523–D531 (2023).

42. Martin, F. J. et al. Ensembl 2023. Nucleic Acids Res. 51, D933–D941 (2023).

43. Brown, G. R. et al. Gene: a gene-centered information resource at NCBI. Nucleic Acids Res. 43, D36–D42 (2015).

